# Consequences of Heterogeneity in Aging: Parental Age at Death Predicts Midlife All-Cause Mortality and Hospitalization in a Swedish National Birth Cohort

**DOI:** 10.1101/2023.07.13.23292617

**Authors:** Anna Thalén, Anders Ledberg

## Abstract

**Background:** The processes that underlie aging may advance at different rates in different individuals and an advanced biological age, relative to the chronological age, is associated with increased risk of disease and death. Here we set out to quantify the extent to which heterogeneous aging shapes health outcomes in midlife by following a Swedish birth-cohort and using parental age at death as a proxy for biological age in the offspring.

**Methods:** We followed a nationwide Swedish birth cohort (N= 89 688) between the ages of 39 to 66 with respect to hospitalizations and death. Cox regressions were used to estimate the association, in the offspring, between parental age at death and all-cause mortality, as well as hospitalization for conditions belonging to one of 10 different ICD-10 chapters.

**Results:** Longer parental lifespan was consistently associated with reduced risks of hospitalization and all-cause mortality. Differences in risk were mostly evident before the age of 50. Each additional decade of parental survival decreased the risk of offspring all-cause mortality by 22% and risks of hospitalizations by 9 to 20 percent across the ten diseases categories considered. The number of deaths and hospitalizations attributable to having parents not living until old age were 1500 (22%) and 11000 (11%) respectively.

**Conclusions:** Our findings highlight that increased parental lifespan is consistently associated with health benefits in the offspring across multiple outcomes and suggests that heterogeneous aging processes have clinical implications already in midlife.

## 1. Introduction

Age is a major risk factor for morbidity and mortality (Niccoli & Partridge, 2012) and interventions aiming at slowing down the pace of aging hold great promise of prolonging both the health span (period of life spend without major illnesses) and the lifespan (age at death) (Kennedy et al., 2014; Partridge et al., 2020). Aging is not a unitary process and theories of aging have postulated a number of different, but interdependent, mechanistic processes through which aging manifests (Kirkwood, 2005; López-Otín et al., 2013, 2023; Strehler, 1977). How these aging-related processes can be controlled is just beginning to be understood (López-Otín et al., 2023), but it is clear that they may evolve differently both within and between individuals; thus generating the heterogeneous phenotype of aging. Indeed, at the population level, some individuals age faster than others, as evidenced by the substantial variability in various measures of aging among people of the same chronological age (Belsky et al., 2015; Lowsky et al., 2014) as well as in the large variability of age at death (Edwards & Tuljapurkar, 2005). The concept of “biological age” has been used to capture the state of aging and to differentiate it from chronological age; a biological age that is higher than the chronological age implies increased risk of disease and death. Given the multifaceted nature of the aging process, it is not surprising that biological age is co-determined by a large number of factors. On the one hand, there are multiple genetic and environmental determinants of biological age (Tian et al., 2023), and, on the other hand, biological age is also dependent on events and behaviors occurring throughout the life course (Kuh et al., 2014; Poganik et al., 2023; Sugden et al., 2023). While age-related deterioration may be most obvious at older ages, variability in measures of aging have been shown in studies also on people of working-ages (Belsky et al., 2015, 2022; Moffitt et al., 2017). However, the extent to which morbidity and mortality in the working-age population can be accounted for by variability in biological age is not fully known, but could provide important information for a fuller understanding of disease etiologies. Moreover, such knowledge could be used in shaping more equitable healthcare policies (Elliott et al., 2021), and for estimating the potential benefits of anti-aging interventions. In this work we aim at quantifying health consequences of heterogeneous aging in a Swedish birth-cohort followed between ages 39 and 66 years.

Since aging and the risk of age-related diseases likely are intrinsically connected, i.e., generated by the same underlying mechanisms (Franceschi et al., 2018; Gladyshev & Gladyshev, 2016), probing the effects of differential aging on disease and death in a population is somewhat challenging. Indeed, that age-related diseases are related to age is tautological, and associations between biomarkers of aging and incidence of age-related diseases and death will be confounded if the biomarkers themselves are caused by the same processes that cause disease and death. To bypass this potential source of confounding, here we instead draw upon the known inheritance of lifespans and use parental age at death as a proxy for biological age in the offspring (Joshi et al., 2016; Timmers et al., 2019). Indeed, it has been long observed that lifespan of parents and their offspring are associated (Pearl, 1922). Some of this inheritance is genetic (Herskind et al., 1996; Hjelmborg et al., 2006), and some is transmitted through other mechanisms (e.g. cultural and economic inheritance). Health outcomes associated with parental lifespans have previously been investigated mainly by comparing offspring from exceptionally long-lived families with controls from the general population, and such studies have consistently shown that risk of disease, as well as risk of death, is lower in probands from long lived families (Ash et al., 2015; Christensen et al., 2020; Dutta et al., 2013; Engberg et al., 2009; Florez et al., 2011; Terry et al., 2004; Westendorp et al., 2009). Here we extend previous work by following a national birth-cohort of Swedish people of working age (from 39 to 66 years of age) and quantify the extent to which mortality, and disease incidence for the 10 most common disease categories, among cohort members, can be accounted for by parental age at death.

## 2. Data & Methods

### 2.1 Data Materials

All data used here come from the longitudinal research programme Reproduction of Inequality through Linked Lives (RELINK-53), which contains information on all persons born in 1953 that lived in Sweden in 1960, 1965 and/or 1968 (N=109 034, referred to as index persons in the following), and their parents (N=213 881) (Almquist et al., 2020). The particular sample and variables used are detailed below.

Personal identification numbers were used to create individual-level linkages from index persons to registries belonging to Sweden’s official statistics. Information about in-patient care and mortality was retrieved from the National Patient Registry (Patientregistret) and Causes of Death Registry (Dödsorsaksregistret) respectively, both administered by the National Board of Health and Welfare. From Statistics Sweden (SCB) data on linkages to biological parents was retrieved from the Multigeneration registry, data on migration and civil status from the Total Population Registry and data on educational attainment from the Longitudinal Integrated Database for Health Insurance and Labor Market Studies.

Linkages between index persons and parents had a lower coverage for index persons that died between 1966 and 1991, and for that reason start of follow-up was set to the first day of the index person’s birth month in 1992 (we did not have access to the exact day of birth). Data on migration were available for the entire cohort up until 2016. For year 2017-2019 migration status was imputed from information on civil status. Full-coverage in-patient and mortality data was available until end of December 2019 which resulted in a 27 to 28 years follow-up period between the cohort member’s 39th birthday in 1992 and the 31st of December 2019. Parents and index persons that emigrated from Sweden before 1992 and did not return before start of follow-up were excluded. Index persons who emigrated after 1992 were followed until their date of emigration in the survival analyses. Moreover, index persons with missing parental information were removed as were those who died before the start of follow-up and those with missing educational information. Parental survival above the age of 55 was set as the lower threshold for parental attained age and index persons with parents who died before age 55 were excluded (n= 9955). This was done to separate the potential difference between premature deaths and deaths that could be attributed to the normal aging process, and to minimize the possible effects of early parental bereavement. Including these index persons with at least one parent who died before age 55 in the analysis did not change results qualitatively (not shown). Parents who, according to the official records, lived to be more than 115 years old were excluded as these cases are likely to reflect persons who de facto have emigrated from Sweden but still appear as residents in the registers (n=21) (Statistics Sweden, 2015). The final sample consisted of 89 688 index persons and their 179 376 parents.

### 2.2 Ethics statement

The research reported here was approved by the Swedish Ethical Review Authority under the RELINK-53 project (no.2017/34–31/5; 2017/684–32).

### 2.3 Procedures

In-patient care records were obtained from the Swedish National Patient Registry and were used as indicators of disease among the index persons. The National Patient Registry uses the Swedish version of the World Health Organization’s International Classification of Diseases (ICD) framework, with version 9 of the ICD in use between 1987 and 1996. After 1996, diagnoses related to in-patient care were coded according to ICD-10. In-patient care visits between 1992 and 1996 were converted from ICD-9 to ICD-10 codes.

To classify in-patient care records into disease categories, the chapter structure of the ICD-10 was used. ICD codes belonging to the 10 most common ICD-10 chapters in this birth cohort were included in the study (see Table 2). We retrieved information on all-cause mortality for both index persons and parents from the National Cause of Death registry. For the parents our main interest was in age at death; however, some parents were still alive (n=17 221, 10%) and we therefore used “parental attained age” to describe the age of the parent. Parental attained age was taken as age at death, or in cases where parents were still alive, their age on the 21st of May 2021 (which was the last date for which we had mortality data). The minimum age of parents who were still alive at this date was 80 years. Index persons’ vital statuses were followed until the end of December 2019 as this was the last date for which we had data on both morbidity and mortality. Thus, the 31th of December 2019 is the last possible day of follow-up for the index persons. Most results are presented as a function of the average attained parental age, i.e., the arithmetic mean of parents’ attained ages. For visualization, average attained age was divided into quartiles: Quartile 1: 55.8-76.5 years, Quartile 2: 76.5-82 years, Quartile 3: 82-86.9 years, Quartile 4: 86.9-108 years.

### 2.4 Statistical Analysis

#### 2.4.1 All-cause mortality

Cox proportional hazard models were used to quantify the association between average parental attained age and the hazard of death. The “adjusted model” was stratified by educational attainment at age 37, parents’ birth years (categorical variable with four level), and sex. Educational attainment reflected the cohort members level of education in the year 1990 and was divided into three categories: elementary school (n=21 216), upper secondary school (n=41 260) and post-secondary education (n=27 212). To visualize the results, cumulative hazard curves were estimated for men and women and the four quartiles of average parental attained age separately (see Figure 1).

**Figure 1.**
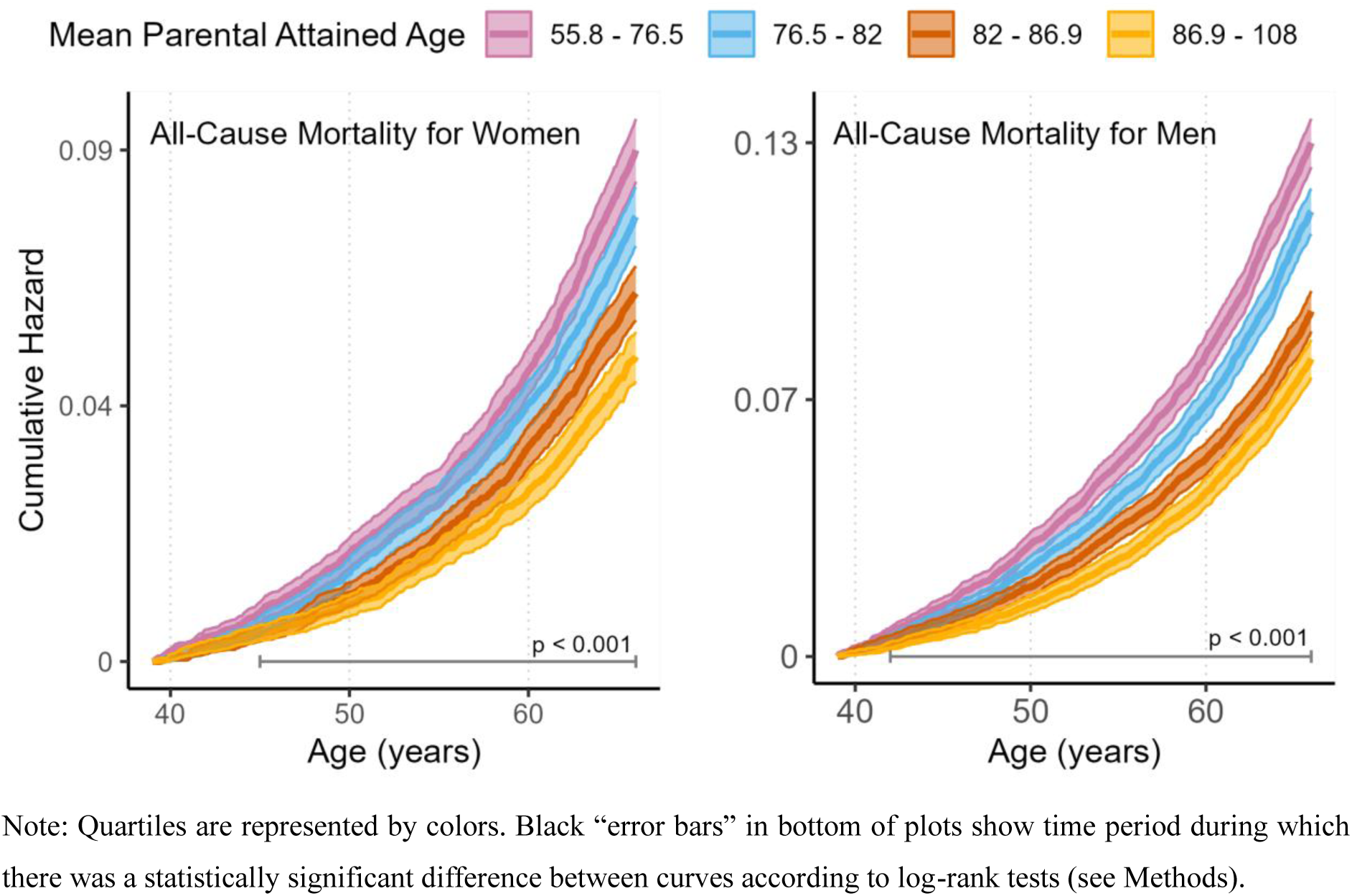
Cumulative risk of index persons’ all-cause mortality (1-survival) stratified by quartiles of average parental attained age

#### 2.4.2 Hospitalizations

To estimate the risk of hospitalization we adopted a competing risks framework where, for each included ICD-10 chapter separately, follow-up was from the date of the index person’s 39th birthday until: emigration, death, diagnosis of interest, or end of follow-up, whichever came first. Hospitalization, death and emigration were considered competing events. Cox proportional hazard models were used to estimate the effect of average parental attained age on the hazard of hospitalization and adjusted models were stratified by the same variables as all-cause mortality. For visualization, the risks of hospitalization are illustrated by cumulative hazards curves, shown as functions of average parental attained age divided in quartiles for both genders combined.

#### 2.4.3 Onsets of risk differences

To identify the age at which the association between parental age at death and risk of all-cause mortality and hospitalization first became noticeable, we recursively applied log-rank tests, testing for risk difference between the four age-quartiles, to data from each ICD-10 chapter and mortality separately. Thus, the first test, for a given outcome, included the complete follow-up, i.e. until last of December 2019. If this test was significant (at *p* < 0.001), a new test was made based on follow-up until last of December 2018. This sequence was then repeated until a non significant test was obtained. The last age for which a significant test was obtained was taken as the onset of the risk difference. In Figures 1 and 2, the ranges of ages with a significant log rank test are indicated by a horizontal line.

**Figure 2.**
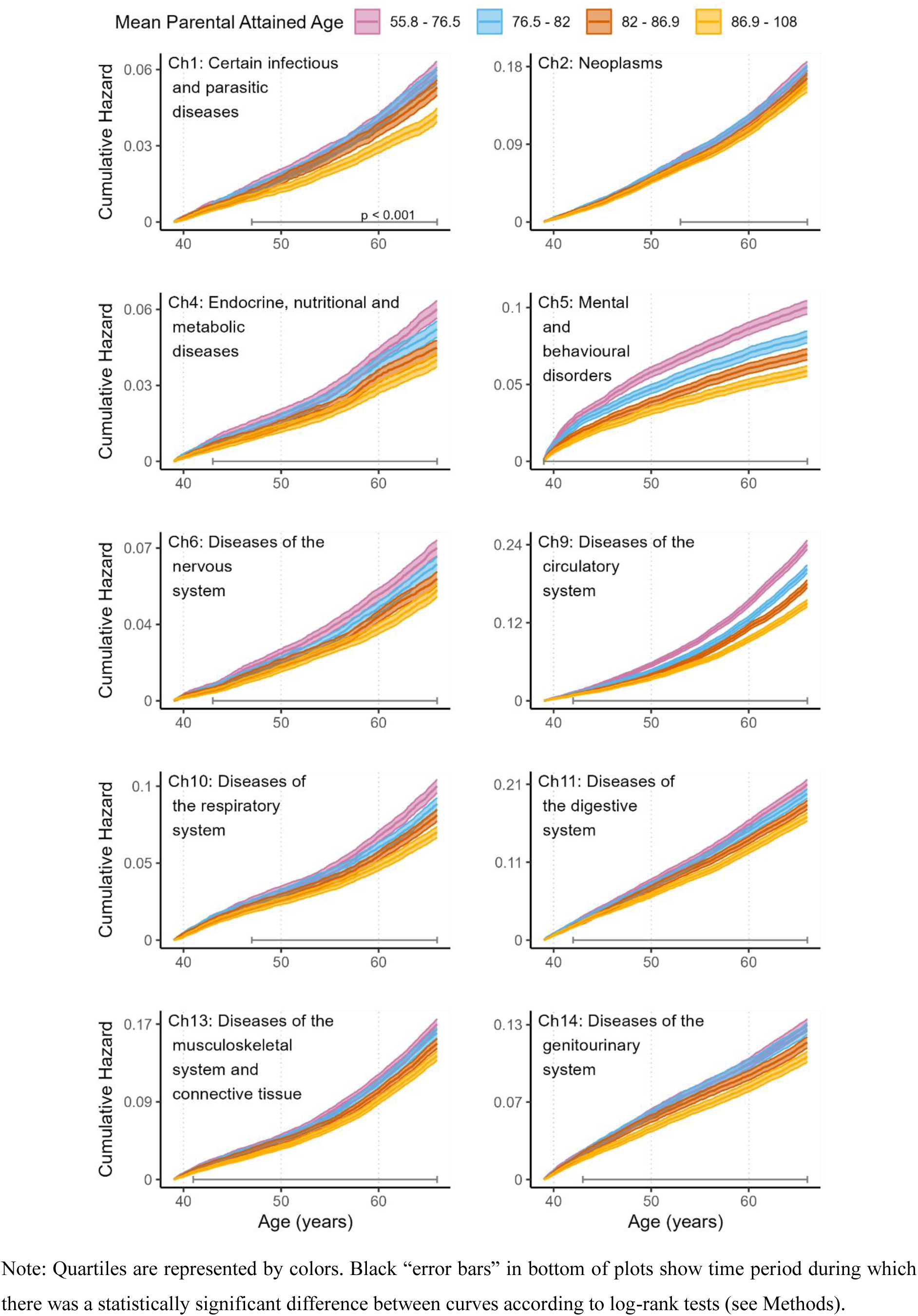
Cumulative risk of index persons’ hospitalization stratified by quartiles of average parental attained age.

#### 2.4.4 Estimating the impact of a hypothetical intervention

The results from the Cox models are expressed in terms of relative hazards rates, and to describe the magnitude of the association in, perhaps, more interpretable terms, we generated non-parametric predictions of the expected number of deaths and hospital admissions under the hypothetical intervention that everyone had parents who lived into the fourth age quartile. The predictions were generated by assuming that the association between parental attained age and risk of all-cause mortality and hospitalization is a causal one, after adjusting for educational attainment and sex. Under this assumption we can estimate the (hypothetical) effect size by resampling from the observed data. For example, if the group of men with elementary school education and average parental attained age in the first quartile, consisted of *m* persons, then, the expected number of deaths in this group, given that they would all have had average parental attained age in the fourth quartile, can be estimated by sampling *m* persons (with replacement) from the group of men with elementary school education and parental attained age in the fourth quartile. To reduce sampling errors, we used the average of 20 resamples. For mortality, the predictions from this procedure correspond closely to predictions from the corresponding Cox model. For hospitalizations, predictions based on the Cox model are harder to make without making additional assumptions. Note that we are not claiming that the effects we report in fact *are* causal; what we do is to provide an answer to the hypothetical question: “if the associations we observe would be causal, what would the effect be of making all people in the data have parents who lived into the fourth quartile?”.

#### 2.4.5 Software

All computations were made in R (R Core Team, 2021). Survival models were estimated using the survival package for R (Therneau, 2021) and plots were made using the ggplot2 package (Wickham, 2016). Necessary data and R-code to reproduce the figures and tables will be made available upon publication.

## 3. Results

The main results reported are based on data from 89 688 people, all born in 1953, who were followed for 27 to 28 years when they were between the ages of 39 to 66. The main independent variable is the average of the parents’ attained age. Analyses where the attained ages of mothers and fathers were entered separately gave similar results (not shown) and for conciseness we use the average attained age here.

### 3.1 All-cause mortality

Nine percent of the index persons died during follow-up, 4971 of which were men, and 3129 women. The risk of dying was associated with parental attained age (Figure 1). Indeed, the cumulative hazard curves show an orderly decrease in the risk of death with increasing parental attained age. As indicated by the horizontal bars, the risks of death differ between the quartiles from relatively young ages: in the early forties for men and mid-forties for women.

To further quantify the association between risk of death and parental attained age, regression models were fit to the data (see Methods), and are reported in Table 1. Increasing average parental attained age by a decade corresponded to a 25 percent reduction in hazard rates in the unadjusted models. Adjusting for educational attainment, sex, and parent’s birth years had only minor effects on hazard ratios (Table 1).

**Table 1.** Adjusted and unadjusted estimates from Cox regression on all-cause mortality on average parental attained age

| Adjusted Model |  |  | Unadjusted Model |  |  |
| --- | --- | --- | --- | --- | --- |
| HR | 95% CI | p-value | HR | 95%CI | p-value |
| 0.78 | 0.76 – 0.81 | <2e-16 | 0.75 | 0.72 – 0.77 | <2e-16 |

Note. Hazard ratios (HRs) are in units of per decade of average parental attained age. The adjusted model was stratified by sex, educational attainment and parental years of birth (see method).

**Table 2.** Number of first-time hospitalizations per ICD-10 chapter during the follow-up period by quartiles of average parental attained age.

| ICD-10 Chapter | Mean Parental Attained Age |  |  |  |  |  |  |  |  |
| --- | --- | --- | --- | --- | --- | --- | --- | --- | --- |
|  | Q1 |  | Q2 |  | Q3 |  | Q4 |  | Total |
|  | 55.8 – 76.5 |  | 76.5 – 82 |  | 82 – 86.9 |  | 86.9 – 108 |  |  |
|  | n | % | n | % | n | % | n | % | n |
| Index persons | 22414 | 25% | 22444 | 25% | 22418 | 25% | 22412 | 25% | 89688 |
| 1: Infectious & parasitic (A00-B99) | 1355 | 6.0% | 1319 | 5.9% | 1226 | 5.5% | 983 | 4.4% | 4883 |
| 2: Neoplasms (C00-D48) | 3644 | 16.3% | 3611 | 16.1% | 3454 | 15.4% | 3256 | 15.7% | 13965 |
| 4: Endocrine, nutritional & metabolic (E00-E90) | 1204 | 5.4% | 1054 | 4.7% | 919 | 4.1% | 823 | 3.7% | 4000 |
| 5: Mental & Behavioral (F00-F99) | 2033 | 9.1% | 1674 | 7.5% | 1441 | 6.4% | 1234 | 5.5% | 6382 |
| 6: Nervous system (G00-G99) | 1483 | 6.6% | 1335 | 5.9% | 1212 | 5.4% | 1098 | 4.9% | 5128 |
| 9: Circulatory system (I00-I99) | 4714 | 21.0% | 4102 | 18.3% | 3705 | 16.5% | 3146 | 14.0% | 15667 |
| 10: Respiratory system (J00-J99) | 2116 | 9.4% | 1894 | 8.4% | 1740 | 7.8% | 1523 | 6.8% | 7273 |
| 11: Digestive system (K00-K93) | 4173 | 18.6% | 3980 | 17.7% | 3711 | 16.6% | 3425 | 15.3% | 15289 |
| 13: Musculoskeletal & conn. tissue (M00-M99) | 3472 | 15.5% | 3303 | 14.7% | 3125 | 13.9% | 2884 | 12.9% | 12784 |
| 14: Genitourinary system (N00-N99) | 2743 | 12.2% | 2676 | 11.9% | 2461 | 11.0% | 2240 | 10.0% | 10120 |

Note. Parental age distribution in each quartile (Q): Q1: 55.8-76.5; Q2: 76.5-82; Q3: 82-86.9; Q4: 86.9-108. We have abbreviated the names of the corresponding ICD- chapters. ICD10-codes used for each included chapter are shown in parenthesis. ICD9 codes used were the following: Infectious: 001-139, Neoplasms: 140-239, Endocrine:240-279, Mental & Behavioral:290-319, Nervous system: 320-389, Circulatory system: 390-459, Respiratory system: 460-519, Digestive system: 520-579, Genitourinary system: 580-629, Musculoskeletal system: 710-739.

### 3.2 Hospitalization

During follow-up, there were in total 95 491 records of in-patient care under a diagnosis corresponding to one of the 10 ICD-10 chapters we investigated. This corresponded to care occasions for 53 052 unique individuals, 27 071 of which were men, and 25 981 women; implying that approximately 59 percent of both men and women were hospitalized at least once during follow-up. Note that this also means that many were hospitalized more than once, i.e., for diagnoses from more than one chapter, and that 41 percent of index persons were not hospitalized with any diagnosis belonging to the ICD-10 chapters we investigated here. The number of first-time hospitalizations per ICD-10 chapter and by parental attained age is presented in Table 2. For most diagnoses, the fraction of in-patient care occasions was lower among index persons whose parents belonged to the fourth quartile of parental attained age.

To study the risk of hospitalization in more detail we did competing risk analyses for the 10 ICD chapters separately. Figure 2 illustrates how the risk of hospitalization varied with parental attained age, and show parental-age-dependent risks for all included ICD chapters. Indeed, increasing parental attained age is consistently associated with a decreased risk of hospitalization. This is particularly clear for chapter 9 (diseases of the circulatory system) and chapter 5 (mental and behavioral disorders). Figure 2 also shows that the risk curves start to diverge well before age 50 for all disease categories except for chapter 2 (Neoplasms).

To further quantify how the risk of hospitalization varied with parental attained age, regression models were fit to the data (Methods), and results are shown in Table 3 and Figure 3. These results confirm the impression from the plots: for all 10 ICD-10 chapters the hazard ratios were clearly below 1. For 8 out of 10 included chapters, one additional decade of parental survival decreased the hazard ratio by 10% or more. Adjusting for sex, education, and parental year of birth reduced the hazard rates slightly.

**Figure 3.**
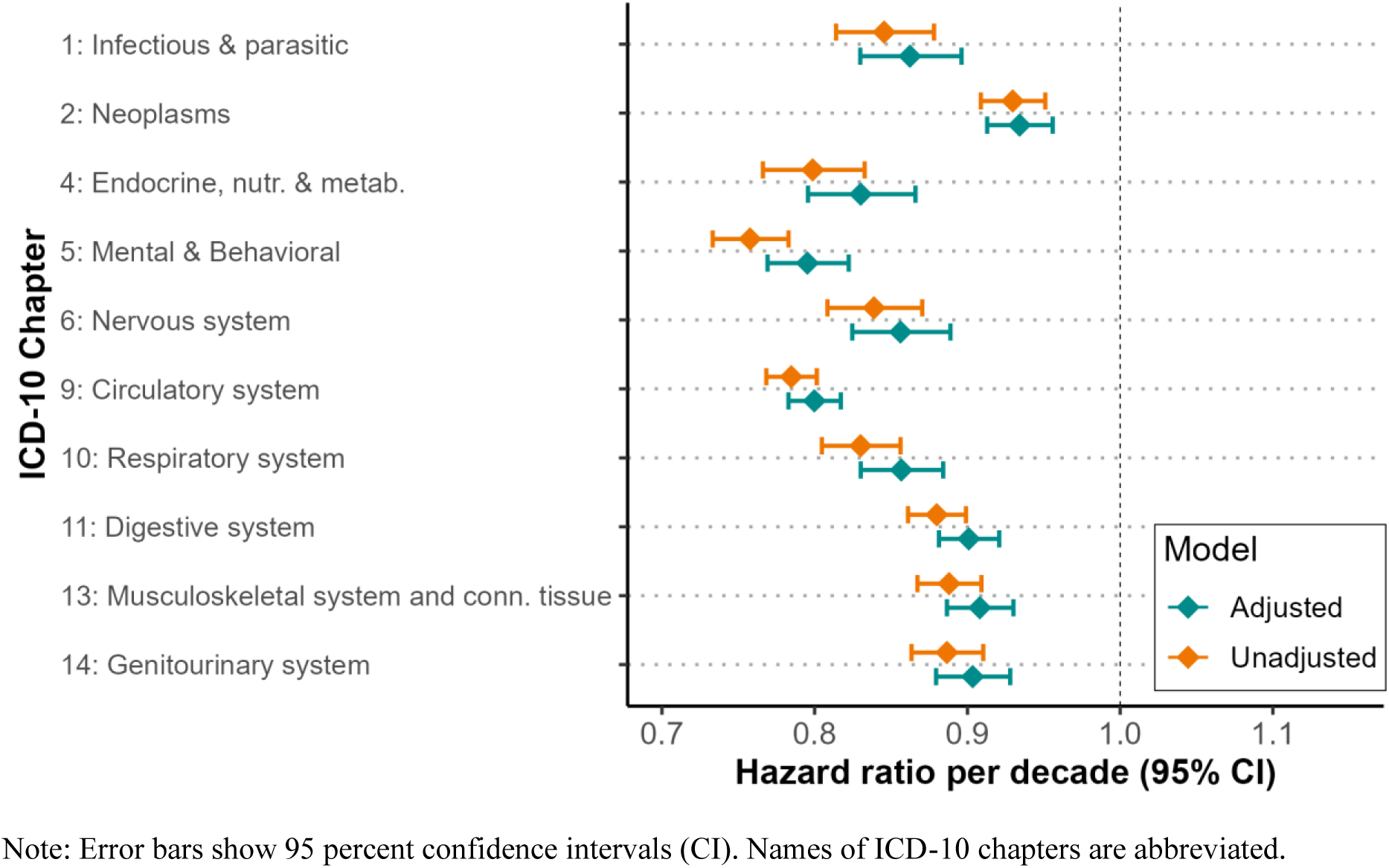
Forest plot of adjusted and unadjusted hazard ratios per decade for average parental attained age for each ICD-10 chapter

**Table 3.** Adjusted and unadjusted estimates from Cox regression for each ICD10-chapter on average parental attained age

| ICD-10 chapter | Adjusted |  |  | Unadjusted |  |  |
| --- | --- | --- | --- | --- | --- | --- |
|  | HR | 95%CI | p-value | HR | 95%CI | p-value |
| 1: Infectious & parasitic | 0.86 | (0.83-0.90) | 4e-14 | 0.85 (0.81-0.88) |  | 4e-18 |
| 2: Neoplasms | 0.93 | (0.91-0.96) | 6e-09 | 0.93 (0.91-0.95) |  | 3e-10 |
| 4: Endocrine, nutritional & metabolic | 0.83 | (0.80-0.87) | 6e-18 | 0.80 (0.77-0.83) |  | 3e-26 |
| 5: Mental & Behavioral | 0.80 | (0.77-0.82) | 2e-41 | 0.76 (0.73-0.78) |  | 8e-62 |
| 6: Nervous system | 0.86 | (0.82-0.89) | 5e-16 | 0.84 (0.81-0.87) |  | 1e-20 |
| 9: Circulatory system | 0.80 | (0.78-0.82) | 2e-93 | 0.78 (0.77-0.80) |  | 5e-113 |
| 10: Respiratory system | 0.86 | (0.83-0.88) | 6e-22 | 0.83 (0.80-0.86) |  | 5e-32 |
| 11: Digestive system | 0.90 | (0.88-0.92) | 9e-21 | 0.88 (0.86-0.90) |  | 3e-31 |
| 13: Musculoskeletal & conn.tissue | 0.91 | (0.89-0.93) | 3e-15 | 0.89 (0.87-0.91) |  | 5e-23 |
| 14: Genitourinary | 0.90 | (0.88-0.93) | 1e-13 | 0.89 (0.86-0.91) |  | 5e-19 |

Note: Hazards ratios (HRs) are in units per decade for average parental attained age. Adjusted estimates are from models stratified by gender, education and parent's birth years. P-values are rounded. Note that 1e-3 = 0.0001.

### 3.3 Impact

To better interpret the magnitude of the associations between parental age at death and all cause mortality and morbidity among offspring, we next asked what the (counterfactual) reduction in number of deaths, and hospitalizations, would have been if everyone in the cohort would have had parents who lived into the oldest age quartile (86.9-108 years). This showed that on average 22% of deaths in the population (corresponding to about 1500 cases) would have been avoided under this hypothetical intervention. For hospitalizations, the corresponding number was 11% (corresponding to a reduction of about 11 000 cases of first-time hospitalizations).

### 3.4 Sensitivity analyses

Sensitivity analyses for the Cox regression models were made which included the full range of parental attained ages that were available in the dataset (ie. the lower threshold of 55 for attained age was removed). Furthermore, a sensitivity analysis which excluded parents who were still alive at the end of follow-up (n= 17 221) was made to ensure that the findings were not driven by the parents who were still alive. The results from these analyses show similar results as those presented above (not shown).

## 4. Discussion

We have investigated how variability in parental attained age, as a proxy of biological aging of the offspring, is related to the manifestation of disease and all-cause mortality in a Swedish nation-wide birth-cohort of people followed between ages 39-66 years. We found that increased parental attained age was consistently linked to reduced risks of being hospitalized for conditions belonging to the 10 most frequent ICD chapters, as well as reduced risk of all cause mortality during the follow-up. When index persons were divided into quartiles based on their parents’ attained ages, differences in risks of hospitalization and death became apparent well before the index persons turned 50 years of age. To quantify the magnitude of the association we showed that a hypothetical intervention, letting everyone in the cohort have parents who lived into the fourth age-quartile, led to a reduction of deaths with 22% and disease incidence with 11%. Arguably, these are quite sizable effects, highlighting the potential power of anti-aging strategies for improving public health.

Some previous studies have investigated the association between parental age at death and health outcomes in offspring of a similar age range as we did here. Dutta et al. (2013) studied a US sample of 6000 persons, aged 51-61 at start of follow-up, and found inverse associations between parental age at death and risk of death as well as incidence of cancer, diabetes, heart disease, and stroke in the offspring. The magnitude of the associations they reported were similar to the ones we found here. Atkins et al. (2016) followed 186 000 participants in the UK Biobank, who were 55-73 years of age at start of follow-up. They found that parents’ ages of death were inversely related to a number of cardiovascular outcomes, with associations of similar magnitude to the ones we report here. Christensen et al. (2020) contrasted the risk of death, and disease incidence, in children and grandchildren from long-lived Danish families with a matched sample from the general (Danish) population. They used hospitalizations for a large number of different conditions, divided into ICD-chapters, as outcomes, and found that children, and to a lesser extent, grandchildren, from long-lived families had a markedly reduced risk for mortality as well as risk for hospitalization for a majority of the ICD-chapters. The age range of the children (offspring) in the study of Christensen et al. (2020) was similar to ours, albeit with a younger minimum age. Interestingly, the grandchildren generation were aged 0-49 during follow-up, and their risk of all-cause mortality as well risk of hospitalization for conditions belonging to a number of ICD-10 chapters, was reduced compared to the control group. This is in line with our results showing that differences in risks of death and disease incidence appeared before age 50 for all conditions except neoplasms. Our study extends the results from these previous studies in several ways. Firstly, by following a (almost) complete national birth cohort, we make certain that the magnitudes of the reported associations are representative for the population. Secondly, by following a large sample, all born the same year, we are able to better follow the development of the risk differences as they change as a function of age (Figures 1 and 2). Thirdly, we show that the risk-modulation by parental attained age is not exclusively due to some special features of long-lived families but rather seem to be a graded effect more widely present in the population.

We argue that parental attained age (or age at death if available) might be a useful proxy for biological age in the offspring. Previous studies have shown that part of the inter-generational association in lifespan is genetic (Herskind et al., 1996; Hjelmborg et al., 2006; Kaplanis et al., 2018; Ruby et al., 2018), and genome-wide association studies (GWAS) have identified several loci associated with lifespan (Joshi et al., 2016; Timmers et al., 2019). This implies that offspring inherit a genetically transmitted propensity for living longer (or shorter) from their parents, and this can reasonably be interpreted as an inheritance of pace of aging. The “frailty model” developed by James Vaupel and co-workers provide a simple instantiation of such mechanisms (Vaupel et al., 1979). According to this model, heterogeneity of lifespans is partly caused by a “frailty factor” which acts multiplicatively on the mortality rates. Thus, if the offspring inherit this “frailty” from their ancestors, they would also inherit a longer (or shorter) lifespan. Vaupel (1988) showed that even a perfect inheritance of frailty may only lead to a modest inheritance of lifespan; in accordance with available data. We note that in our study, the mortality and morbidity risks seemingly depended on parental attained age in a multiplicative way; in good agreement with predictions from the frailty model. The interpretation of parental attained age as an indicator of biological age in offspring is further supported by the results we report here showing a consistent association between parental age at death and risk of most major diseases, as well as all-cause mortality. Additional support for this interpretation was provided by Christensen et al. (2020) who showed that inheritance of health- and lifespan in their study did not seem to be disease specific, but rather reflected “fundamentally slower aging”.

While parental attained age likely reflects both genetic and environmental factors influencing biological age, there are certainly other factors, such as individual behaviors and life course exposures that also contribute to variability in biological aging Moreover, the exact age at which someone dies is dependent on many other things in addition to biological age, meaning that parental attained age is at best a noisy indicator of biological age in the offspring. Future work should compare variability in biological aging accounted for by parental attained age to that accounted for by biomarkers of aging (Jylhävä et al., 2017) to find more comprehensive ways to measure biological age.

In conclusion, our study reveals that parental attained age is consistently associated with a substantially reduced morbidity and mortality in offspring of working ages, and that these risk differences are in many cases noticeable already before age 50. These results suggest that a sizable part of the disease burden in midlife can be attributed to an inheritable heterogeneity in the aging process.

## Data Availability

Aggregated data will be made available upon publication.

## Acknowledgements

We would like to thank Can Liu for giving helpful comments on the manuscript. We also acknowledge that preliminary results were reported in the master thesis of Thalén (2022).

